# The association between salivary amylase gene copy number and enzyme activity with type 2 diabetes status

**DOI:** 10.1101/2025.03.31.25324922

**Authors:** Sri Lakshmi Sravani Devarakonda, Jennifer Ren, Angela C. Poole

**Author notes:** Corresponding author (ACP).

## Abstract

The association between salivary amylase gene (*AMY1*) copy number (CN) and metabolic health parameters, such as insulin resistance, has been studied but with conflicting results. The cause of these inconsistencies is multifaceted and confounded by differences in genotyping methods. In this study, we investigated the factors that affect the association between *AMY1* CN, salivary amylase activity (SAA), and type 2 diabetes (T2D) status. We collected up to four saliva samples from two cohorts: individuals with self-reported T2D or prediabetes (n = 18) and individuals who self-reported as being without T2D or prediabetes (controls, n = 178). We genotyped individuals for *AMY1* CN using both quantitative PCR and droplet digital PCR and measured SAA in the saliva samples. We demonstrated that these two commonly used methods give comparable values for *AMY1* CN. We showed a positive association between the time of saliva collection and SAA throughout the day. We also observed that *AMY1* CN and SAA are positively associated in individuals with and without T2D or prediabetes. With increasing *AMY1* CN, SAA was higher for each additional copy of *AMY1* CN in individuals with T2D or prediabetes compared to controls. Our findings support the premise that increased SAA in T2D patients is a compensatory mechanism related to the long-term protective effect of high *AMY1* CN on glucose metabolism.

## Introduction

Copy number (CN) variation affects ∼10% of the human genome [1]. CN variants (CNVs) modulate gene expression [2,3] and, as such, are genetic causes of phenotypic variability and disease risk. However, reliable determination of gene CN is challenging. One well known CNV is *AMY1*, which encodes the salivary amylase enzyme. Amylase is the most abundant enzyme in human saliva and initiates starch degradation in the mouth. In humans, diploid *AMY1* CN ranges from 2 to 20 [4], and this variation is thought to affect metabolic health [5–8]. However, there is conflicting evidence regarding the relationship between *AMY1* CN and metabolic disorders such as obesity and impaired glucose homeostasis [9–13]. These inconsistencies in the associations between *AMY1* CN and metabolic health could be partly attributed to the methodological differences in determining *AMY1* CN.

Studies involving *AMY1* have used various methods to determine CN, including quantitative PCR (qPCR) [8,11,14,15], droplet digital PCR (ddPCR) [5,6,10], fluorescence in situ hybridization (FISH) [9,14], paralogue ratio tests (PRT) [9,16,17], and computational approaches [10]. Research groups using these methods to address the same question have come to different conclusions. For example, using qPCR, one of the first studies addressing the association between *AMY1* CN and BMI reported an inverse relationship [11]. By contrast, because of the correlation between *AMY1* CN and historic dietary patterns, an earlier publication proposed that a higher CN conferred the fitness advantage of weight gain due to more efficient extraction of calories from starchy foods [14]—this supports the premise that *AMY1* CN is positively correlated with BMI. Other research groups, using methods other than qPCR to determine *AMY1* CN, reported no association between *AMY1* CN and BMI [9,10]. This provoked questions regarding the accuracy of qPCR versus other methods for *AMY1* CN determination [18]. The first goal of our study was to compare qPCR and ddPCR, two commonly used laboratory-based methods to determine *AMY1* CN; ddPCR is known for accuracy but qPCR is still sometimes used depending on resources available to research teams.

It has been reported that *AMY1* CN is positively correlated with salivary amylase protein quantity and SAA [19]. However, assuming a consistent correlation between *AMY1* CN and SAA in the context of physiological implications could be misleading because several environmental factors contribute to variability in SAA, including the time of day [19,20]. Prior studies have also reported higher salivary amylase concentration or SAA in people with T2D than in healthy controls, and it has been proposed that these measurements could be used as diagnostic biomarkers for T2D [21]. As a second goal in this study, we investigated the association between *AMY1* CN and SAA in healthy individuals compared to those with self-reported T2D or prediabetes and explored the diurnal variation of SAA. Our findings implicate SAA in glucose homeostasis and provide practical insights for future research and health applications involving this gene.

## Materials and methods

### Participant Recruitment and Sample Collection

The human subjects research from which the samples were derived was approved by the Cornell University Institutional Review Board (Approval Number: 1902008575 [starch study] and 1808008178 [T2D microbiome study]). The study population included two cohorts: (1) healthy individuals 18 years or older in the starch study and (2) individuals with a self-reported diagnosis of prediabetes or type 2 diabetes (T2D) 45 years or older in the T2D microbiome study. The demographics of the two cohorts are shown in Table S1. We recruited each cohort from the local community in and around Ithaca, New York, USA. The recruitment periods for the starch study were from August 1, 2019, to December 31, 2019, and from September 2, 2020, to October 2, 2020. The recruitment period for the T2D microbiome study was from October 13, 2019, to March 14, 2020. All participants provided written informed consent before enrollment for both cohorts. To confirm the diagnosis of prediabetes or T2D for each participant in the T2D cohort, we requested a diabetes medication prescription or a doctor’s note. In addition, the participants in the T2D cohort underwent an oral mixed-meal tolerance test to assess glucose dysregulation (Fig S1).

Approximately 5 ml of saliva was collected from the 196 participants for *AMY1* CN determination. In a subset of 94 participants, up to four saliva samples were collected on different days to measure SAA. Before saliva collection, participants were asked to refrain from brushing their teeth for a minimum of six hours and to not consume any food or beverages, including water, for a minimum of 30 minutes. Saliva samples were collected using a non-invasive, self-collection method. Participants were instructed to allow saliva to passively accumulate in their mouth and then expel it into a sterile 50 ml conical tube. These samples were placed on ice immediately after collection, aliquoted within three hours, and stored at -80°C until further analysis.

### DNA extraction

Genomic DNA was extracted from saliva samples using the QIAamp DNA Blood Kit (Qiagen, cat # 51161), the QIAamp DNA Blood Mini Kit (Qiagen, cat # 51104), and the QIAamp DNA Investigator Kit (Qiagen, cat # 56504) following manufacturer protocols.

### *AMY1* copy number determination by quantitative PCR

We performed quantitative PCR (qPCR) using the following primers to amplify the *AMY1* genes, *AMY1A*, *AMY1B*, and *AMY1C*: AMY1-forward: 5’-TGAGAACATTAGGCCACAGCA-3’, AMY1-reverse: 5’-TGGAAATCATCTCAATGACCTCT-3 [22]. We used *EIF2B2* as a reference gene (CN = 2) and the following primers to amplify the reference gene: EIF2B2-forward: 5’-GCTCAAAGTGCTTGAGGACC-3’; EIF2B2-reverse: 5’-CAAAGCCAAACCCAGACAAT-3’. For each gene of interest, the qPCR reaction consisted of 1 μl genomic DNA (5 ng/μl), 0.5 μl of each 10 μM primer (forward and reverse), and 3 μl of PCR grade H_2_O, and 5 μl iTaq™ Universal SYBR® Green Supermix (Catalog # BioRad 1725122), for a total volume of 10 μl per reaction. The qPCR conditions were: initial denaturation at 95 °C for 5 minutes and 40 cycles of 95 °C for 10 seconds and 60 °C for 30 seconds on a Roche LightCycler 480 Real-Time PCR Instrument. A standard curve was made using genomic DNA NA12286 (Coriell Institute; *AMY1* CN = 2). All reactions, including standards and blanks, were performed in quadruplicate. These genomic DNAs from the Coriell institute were used as positive controls on all qPCR plates to assess variability between plates: NA18972, NA12873, NA10472, NA12890, NA10852, NA12043, NA11992, NA12414, NA12340, NA06994, NA12342, NA12286, NA18522, and NA19138.

### *AMY1* copy number determination by droplet digital PCR

Before copy number determination by droplet digital PCR (ddPCR), we digested the genomic DNA with the HaeIII restriction enzyme (New England Biolabs) to separate the tandemly repeated copies of the *AMY1* gene. This mitigates the underestimation of copy numbers in high copy number samples. Each 15 ul restriction digestion reaction consisted of 12 ul genomic DNA at 15-20 ng/ul concentration, the 1.5 ul HaeIII enzyme at 2 U/ul, and 1.5 ul of 10x NEB buffer. DNA was digested for 60 minutes at 37 °C, followed by heat inactivation of the enzyme for 20 minutes at 60 °C.

We determined *AMY1* CN compared to a reference gene *AP3B1* using ddPCR on the Bio-Rad QX100 AutoDG Droplet Digital PCR System. The PCR amplification reaction included 10 ul of 2x ddPCR Supermix for Probes No dUTP (Bio-Rad), 2 ul of restriction enzyme digested genomic DNA at a concentration of 12–16 ng/ul, 0.5 ul of *AMY1* TaqMan assay (Hs07226361_cn, ThermoFisher Scientific), and 0.5 ul of reference gene *AP3B1* assay (dHsaCP1000001, Bio-Rad) for a total reaction volume of 20 ul. The reaction was emulsified with Droplet Generator Oil (Bio-Rad) using the QX100 Droplet Generator (Bio-Rad) per the manufacturer’s instructions. *AMY1* CN was determined using QuantaSoft software version 1.7.4.0917 (Bio-Rad laboratories).

### Salivary amylase activity assay

We used the Salimetrics Salivary Alpha-Amylase Enzymatic Kit (Salimetrics, cat 1-1902) to measure SAA. We performed the assay in triplicate for each saliva sample. We followed the manufacturer protocol except for using 300 ul amylase substrate per reaction instead of 320 ul. The calculations were adjusted according to the change in total assay volume. The triplicate measurements for each participant were averaged to obtain the mean SAA to be used in the analyses.

### Statistical analysis

For all participants, we determined the *AMY1* CN with qPCR and ddPCR in two separate experiments. For statistical analyses comparing qPCR and ddPCR, we calculated the mean CN value for each method by averaging the results of the two separate experiments. For all the analyses examining the association of *AMY1* CN with other parameters, we used the mean CN value obtained from the two ddPCR experiments for each individual.

All statistical analyses were conducted using Rstudio (4.2.2), and a p-value of ≤0.05 was considered statistically significant. We fit linear mixed models to examine how SAA is influenced by *AMY1* CN, the time of saliva sample collection, and T2D status. The response variable, SAA, was log-transformed to meet the assumptions of linear models, and the normality of the model residuals was assessed. We included subject ID as a random effect because multiple samples were collected from each participant at different times. The linear mixed models were conducted with the lme4 package in R [23]. Figures depicting the data points used in the linear models also show the fitted lines of the model-predicted values. We also used linear regression to examine the relationship between *AMY1* CN values determined using qPCR and ddPCR and used intra-class correlation (ICC) to check the consistency between qPCR and ddPCR values. Tables S2– S7 show the R output for each of the models.

## Results

### qPCR and ddPCR estimates for *AMY1* CN

The median *AMY1* CN obtained with qPCR was 7.5 with a range of 2 to 19 copies, and the median *AMY1* CN obtained with ddPCR was 7 with a range of 2 to 20 copies. We compared the *AMY1* CN estimates of the reference Coriell DNAs obtained using qPCR and ddPCR in our lab to the values previously reported in the literature (Table 1). For each Coriell DNA, the standard deviation of all the *AMY1* CN estimates was between 0.20–2.54 with a median standard deviation of 0.73 when we exclude NA10472 and NA10852. The greatest standard deviation in CN estimates was for NA18972, which has the highest *AMY1* CN.

**Table 1.** Comparison of *AMY1* CN estimates of Coriell DNAs determined using PRT, QuicK-mer, qPCR, and ddPCR.

| Coriell DNA | Previous literature |  |  |  | This study |  |
| --- | --- | --- | --- | --- | --- | --- |
|  | PRT [9,24] | Read Depth [9] | qPCR [9] | QuickK-mer [25] | qPCR | ddPCR |
| NA18972 | 18 | 18.6 | 21.61 | 14.2 | 16.35 | 16.16 |
| NA10472 | NA | NA | NA | NA | 6.14 | 6.18 |
| NA10852 | 6 | NA | NA | NA | 6.17 | 5.4 |
| NA11992 | 8.05 | 7.16 | 8.22 | 5.28 | 6.57 | 6.53 |
| NA12340 | 4.78 | 5.04 | 5.13 | 4.63 | 4.9 | 4.66 |
| NA12342 | 2.92 | 2.86 | 1.88 | 2.79 | 3.125 | 2.79 |
| NA18522 | 3.79 | 3.98 | 4.51 | 3.66 | 4.025 | 3.7 |
| NA19138 | 8.19 | 7.97 | 8.63 | 6.42 | 8.05 | 7.81 |
| NA12873 | 7.31 | 12 | 11.37 | 10.44 | 10.29 | 9.93 |
| NA12890 | 8.78 | 9.81 | 10.53 | 8.79 | 10.1 | 9.57 |
| NA12043 | 13.57 | 16.94 | 17.57 | 13.36 | 13.54 | 13.79 |
| NA12414 | 13.61 | 11.54 | 12.95 | 10.01 | 11.18 | 11.38 |
| NA06994 | 8.56 | 8.06 | 8.28 | 7.16 | 7.37 | 7.32 |
| NA12286 | 2.03 | 2.33 | 2.51 | 1.96 | 2.01 | 1.83 |

We used linear regression to examine the association between the CNs determined by qPCR and ddPCR for all 210 samples, which included 196 study participants and the Coriell DNAs. qPCR-estimated *AMY1* CNs were used to predict ddPCR-estimated CNs (Table S2). qPCR measurements were predictive of the ddPCR measurements (β = 0.917, p < 0.001). However, the qPCR-estimated CN values were more dispersed (Fig 1), suggesting that for a given *AMY1* CN, qPCR produced more variable estimates than ddPCR. The variability in qPCR estimates for given ddPCR estimates of *AMY1* CN may indicate lower precision in qPCR, even though the overall variance between the two methods was not statistically different, as shown by the F-test (F= 1.07, df = 209, p = 0.63). To further assess agreement between the two techniques, we calculated the intraclass correlation coefficient (ICC; Table S3). The ICC coefficient was 0.95 (95% CI: 0.93–0.96), indicating a high level of consistency between qPCR and ddPCR in estimating *AMY1* CN.

**Fig 1.**
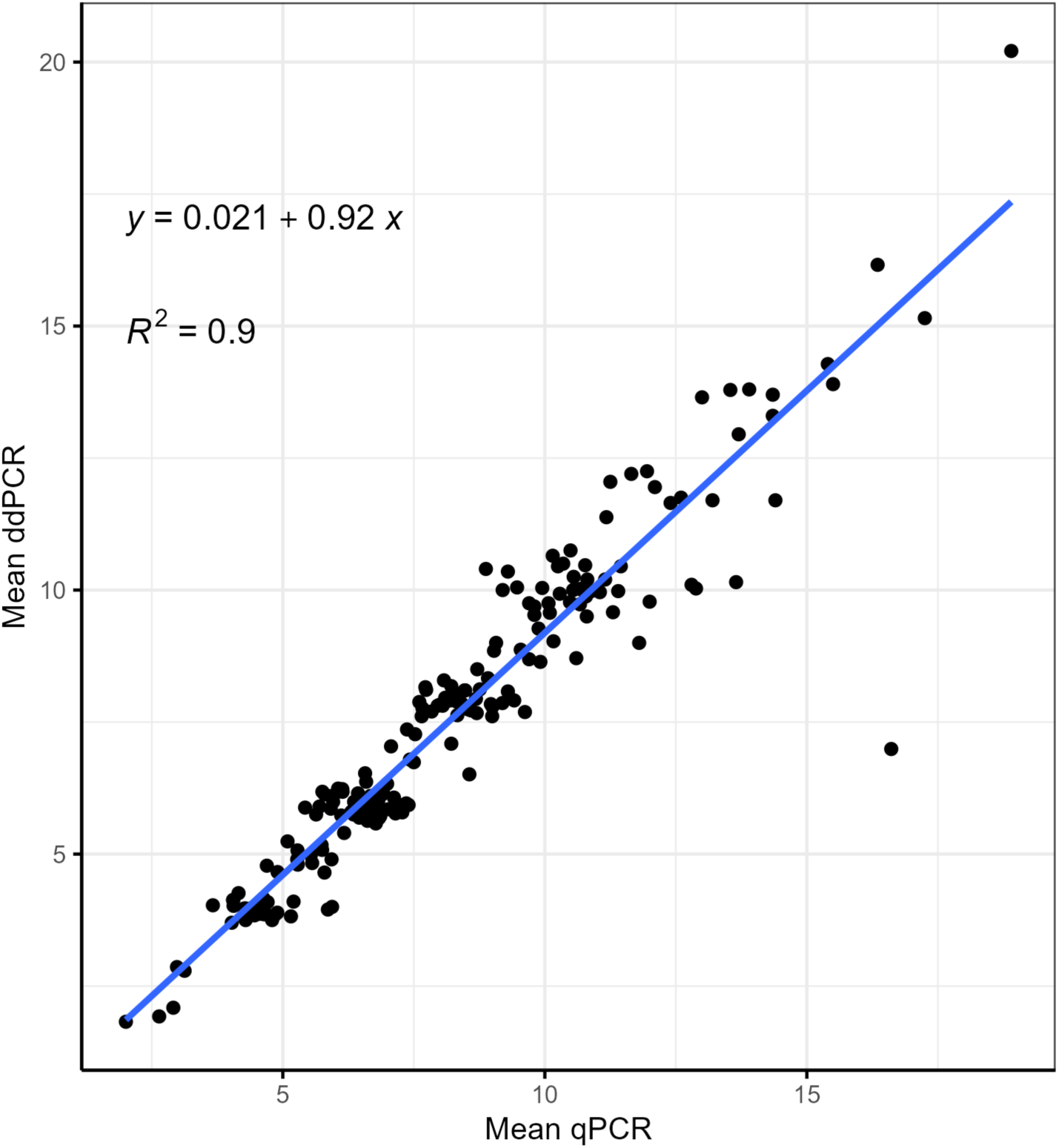
Plot depicting the relationship between *AMY1* CN estimated by qPCR and ddPCR. The values on the x-axis and y-axis are the mean CN of two replicates of qPCR and ddPCR, respectively. For each qPCR replicate, the reactions were run in quadruplicate. The plot shows individual CN estimates with a linear regression line.

### Saliva sample collection time

A linear mixed model was used to examine the diurnal pattern of SAA, with log-transformed SAA as the response variable and time of saliva collection as the predictor (Table S4). Participants were included as a random effect to account for interindividual variability. The results revealed diurnal variation in SAA—SAA increased as the day progressed (n = 263 samples; β = 0.09; p < 0.001). A previous study reported a sharp increase in SAA from morning to noon followed by a slower rate of change throughout the afternoon [26]. To evaluate whether this pattern occurred in our dataset, we ran the model separately for morning (t ≤ 12:00) and afternoon (t > 12:00) samples, to compare the rate of increase in SAA between the two periods. We observed that SAA increased significantly with time in the morning (n = 111 samples; β= 0.18; p = 0.003), and after noon (n = 152 samples; β = 0.085; p = 0.05). For every one hour increase in the time of collection, the expected value of SAA increased by 19% within the samples collected at or before noon. The percent increase was 9% in the samples collected after noon.

To determine whether the difference in the rate of increase in SAA between morning and afternoon was significant, we used a linear mixed model to predict the log of the SAA by the time of saliva collection during the day, the time of day (categorical as morning/afternoon), and their interaction. We also included a random effect of participant to account for repeat measurements from participants (Table S5). There was no significant difference in the rate of increase in SAA with time between morning and afternoon (β= 0.107; p = 0.1) (Fig 2).

**Fig 2.**
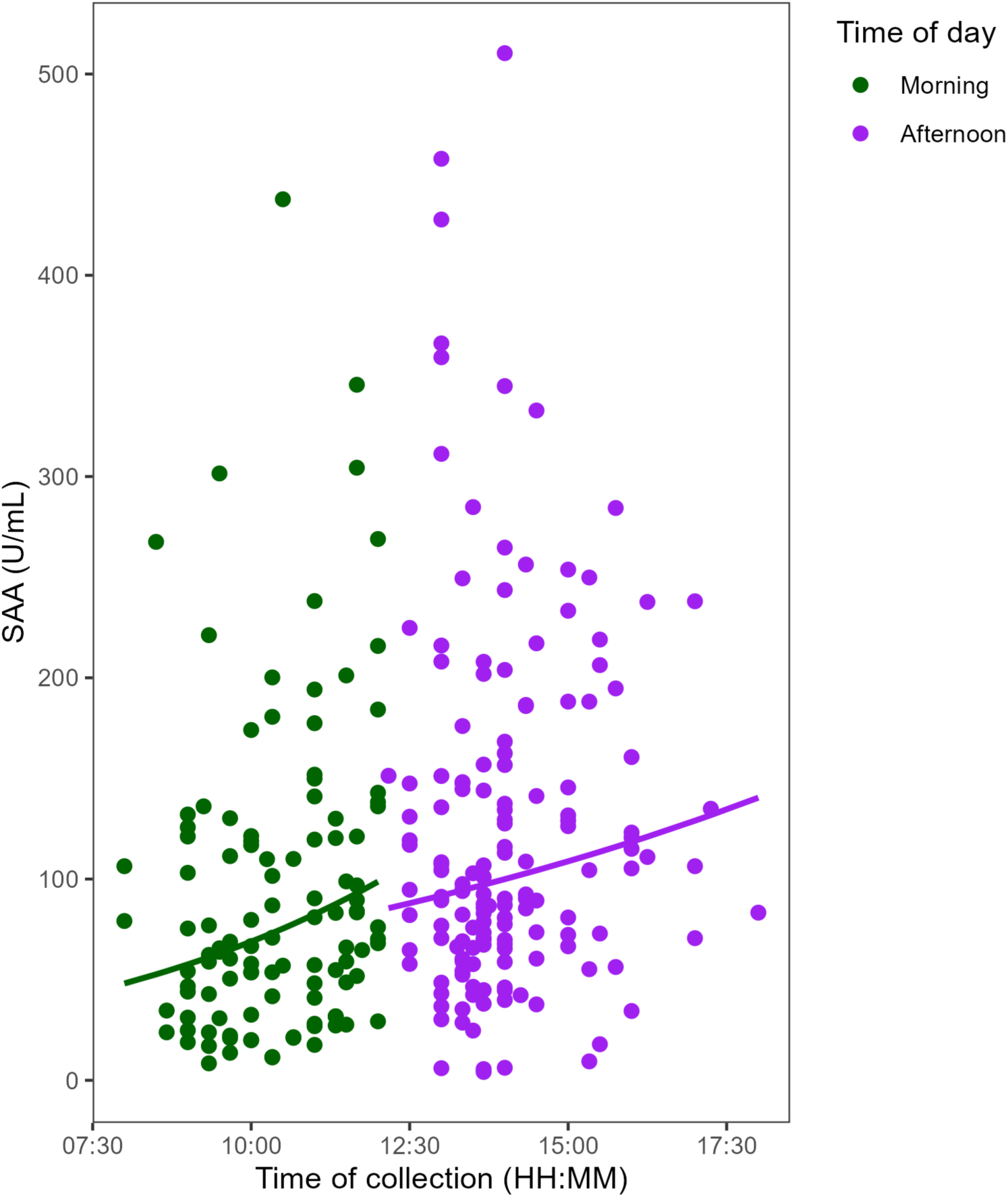
Diurnal increase in salivary amylase activity (SAA). The data points in purple represent samples collected at or before noon (n = 111). The data points in green represent the samples collected after noon (n = 152). The solid lines represent predicted values from the linear mixed model fitted for samples collected at or before noon and after noon. The rate of change of SAA does not significantly differ between morning and afternoon (p = 0.1).

### *AMY1* CN and SAA

We used a linear mixed effects model with log-transformed SAA as the response variable, *AMY1* CN obtained by ddPCR as a fixed effect, and participant as a random effect to investigate the association between *AMY1* CN and SAA (Table S6). We found a significant positive association between *AMY1* CN and SAA (n = 300 samples; β = 0.15; p < 0.001). We assessed model fit using marginal R² and conditional R² in the rsq package [27] in R. *AMY1* CN alone explained 18% of the variance in SAA. There was a high conditional R^2^ (75%) indicating that *AMY1* CN and interindividual differences, due to potential biological or other factors not included in the model, explain most of the variance in SAA.

### T2D/prediabetes status modifies the relationship between ***AMY1* CN and SAA**

To evaluate whether the association between *AMY1* CN and SAA varied in individuals with T2D or prediabetes compared to healthy controls, we fit a linear mixed effects model with the log of SAA as a response variable, and with *AMY1* CN, health status (T2D/prediabetes or healthy control), age, sex, and time of saliva collection as predictors (Table S7). An interaction term between *AMY1* CN and T2D/prediabetes status was included. We excluded individuals with missing data for any variables in the model. The interaction between *AMY1* CN and T2D/prediabetes status was significant (n = 285 samples; β = 0.22; p = 0.006). For each additional copy of *AMY1*, SAA increased by approximately 14% in the control group and 43% in the T2D/prediabetes group. Taken together, with increasing *AMY1* CN, SAA was higher for each additional copy of *AMY1* CN in individuals with T2D or prediabetes compared to controls (Fig 3). These findings suggest that T2D/prediabetes status modifies the relationship between *AMY1* CN and SAA.

**Fig 3.**
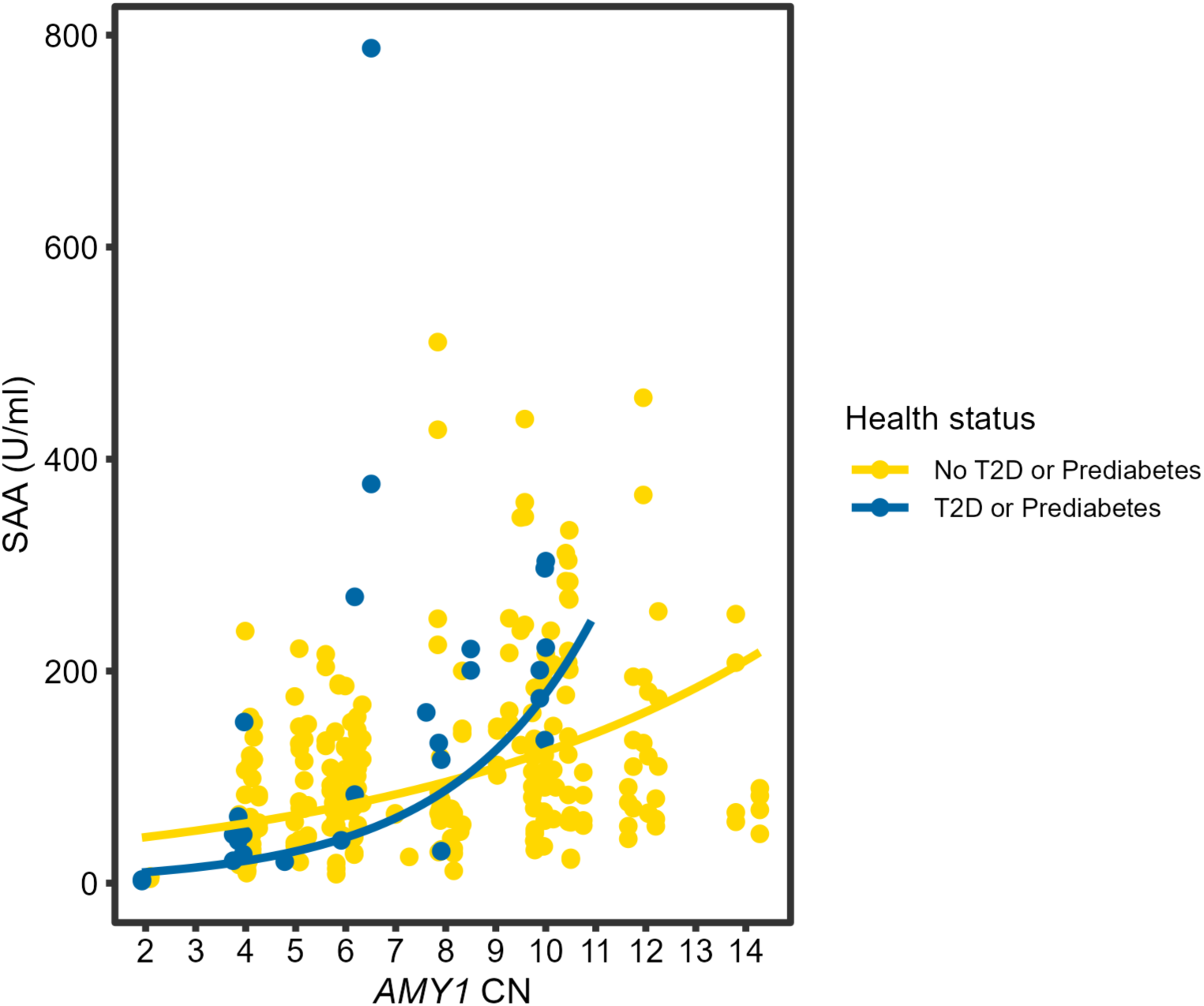
The relationship between *AMY1* CN and SAA based on T2D/prediabetes status. The data points are colored according to the participants’ T2D/prediabetes status with up to four samples collected per individual. For T2D/prediabetes, blue data points n = 27; for healthy controls, yellow data points n = 258. The lines in this figure represent predicted values from the linear mixed model. For the interaction between *AMY1* CN and T2D/prediabetes status, p = 0.006.

## Discussion

The key finding in our study is that T2D/prediabetes status modifies the association between *AMY1* CN and SAA. In addition, we found evidence to support diurnal variations in SAA. We observed a positive correlation between SAA and the time of day at which saliva samples were collected. We also showed that qPCR values can be comparable to ddPCR values when measuring *AMY1* CN.

Whereas *AMY1* CN is a fixed measure, fluctuations in SAA could provide insight about the day-to-day impact of salivary amylase on blood glucose homeostasis. We observed that compared to healthy controls, people with prediabetes or T2D had a greater increase in SAA with an increase in *AMY1* CN. At least four other studies found that T2D patients had elevated SAA compared to controls [28–31]. Our analysis, confirming higher SAA in T2D/prediabetes patients, included samples collected from T2D/prediabetes patients and controls in the morning and after noon. Our statistical model included time of collection as a covariate. Since participants with T2D or prediabetes have aberrant glucose homeostasis, these findings suggest that higher SAA is associated with glucose dysregulation. If we assume that participants with T2D/prediabetes had lower SAA before developing the glucose dysregulation that resulted in their diagnosis, an increase in SAA from one’s previous mean activity may be indicative that disease onset is pending. Therefore, increased SAA could either be contributing to the development of T2D/prediabetes or counteracting glucose dysregulation.

Most studies assessing the long-term effects of salivary amylase dosage and glucose metabolism are reported in terms of *AMY1* CN, as opposed to SAA. Two studies have reported that a higher *AMY1* CN is associated with an increased postprandial glucose response to starches, with both studies also reporting a positive correlation between *AMY1* CN and SAA [7,32]. This result suggests that a higher CN, with presumably higher SAA, would more efficiently liberate sugars from starchy foods to raise blood glucose. When attempting to minimize steep increases in postprandial glucose response to dietary intake for the prevention or management of T2D, a high *AMY1* CN could be a challenge for people with a high starch intake. However, one study reported an inverse association between *AMY1* CN and fasting blood glucose in people with a high starch intake [32]. This suggests there is some protective mechanism stabilizing blood glucose long term in people with high *AMY1* CN. Two studies reported that high SAA is associated with low glucose response to starch in a small number of subjects [33,34]. Mandel and Breslin posited that this is mediated by preabsorptive insulin release, which was observed in high SAA participants [34]. This would support the premise of a protective mechanism with increased SAA being a compensatory effect blunting blood glucose response to starch ingestion. Congruent with the premise that high *AMY1* CN has a protective effect, there are several reports that high *AMY1* CN is associated with decreased insulin resistance and T2D risk [8,12]. Additionally, another large study reported an inverse association between starch intake and T2D in individuals with *AMY1* CN ≥10 [5].

One limitation is that our study was cross-sectional. Since we only measured SAA in each individual during their current disease status—when they were either with or without prediabetes or T2D—we do not know whether the T2D/prediabetes participants had higher SAA throughout their adult lives, or their SAA increased before or after disease onset. A potential limitation of our findings involving SAA and T2D status is the relative sample sizes of the groups—there were 16 individuals with T2D or prediabetes and 74 individuals without T2D or prediabetes included in the analysis. Another potential limitation is the difference in the age range of the healthy cohort (18–59 years) and the T2D/ prediabetes cohort (47–87 years).

However, when we performed a linear regression analysis to investigate the relationship between age and SAA, we found no significant association between the two variables. Furthermore, SAA does not appear to vary as adults age, although the studies addressing this question were cross-sectional [35,36]. A study that would best address the question of whether SAA could be a biomarker for the onset of T2D would be prospective in several populations of different ethnic groups and with multiple risk factors for T2D. This should include *AMY1* CN determination and measurements of SAA and glucose response to starch at multiple time points.

We also demonstrated that qPCR can be a reliable alternative for copy number assessment with comparable accuracy to ddPCR, which has been proposed as the model method for quantifying a target gene copy number [37]. This finding is not consistent with another study that reported discordance between qPCR and ddPCR values for *AMY1* CN in saliva samples [38]. Advantages of ddPCR include sensitivity and no requirement for a standard curve. Conversely, qPCR is less expensive and faster than ddPCR and, thus, more adaptable for high throughput protocols with automation. Another consideration, when using ddPCR for quantifying a target gene CN, is the method poses an inadvertent risk of underestimating CNs when multiple copies of the target gene are linked in tandem [39]. As previous groups have done, we addressed this by initially digesting the genomic DNA using a restriction enzyme. This additional step in the protocol augments the processing time and expense. Thus, qPCR may be preferred over ddPCR for *AMY1* CN estimation if there are a large number of samples being genotyped or there is restricted access to resources.

We pose a question regarding the interchangeability of *AMY1* CN and SAA when studying metabolic health. *AMY1* CN remains stable within an individual through their lifetime. This genetic stability could make it a reliable metric to assess an individual’s metabolic predisposition. Unlike *AMY1* CN, SAA fluctuates and can be affected by several factors, e.g., time of sample collection, caffeine intake, smoking, and stress, in addition to *AMY1* CN [35]. We observed that, although SAA positively correlates with *AMY1* CN, there is diurnal variation. The pattern in SAA is a rapid increase in the morning followed by a slower increase in the afternoon, consistent with previous findings [26]. Further investigation is required to determine the impact of these differences in diurnal patterns on metabolic health. A key implication is that consistent timing of saliva collection across subjects would be ideal when investigating the role of SAA in health and disease.

## Data Availability

All data produced in the present study will be publicly available upon publication of the manuscript in a peer-reviewed journal.

## Acknowledgments

We thank Erika Mudrak from the Cornell Statistical Consulting Unit for assistance with the analyses and Dorothy Kim Superdock for her help with sample collection and processing. We used DNA from human lymphoblastoid cell lines from the NHGRI Sample Repository for Human Genetic Research and the NIGMS Human Genetic Cell Repository of the Coriell Institute for Medical Research. We thank the Biotechnology Resource Center Genomics Facility at the Cornell Institute of Biotechnology for performing ddPCR.

## Notes

### Competing Interest Statement

The authors have declared no competing interest.

### Funding Statement

This study did not receive any funding.

### Author Declarations

The Institutional Review Board of Cornell University gave ethical approval for this work (Approval Number: 1902008575 [starch study] and 1808008178 [T2D microbiome study]).

