## Supplementary material for "The association between salivary amylase gene copy number and enzyme activity with type 2 diabetes status"

**Mixed meal tolerance test in participants with self-reported type 2 diabetes or prediabetes (T2D/prediabetes) to confirm glucose dysregulation.**

A mixed meal tolerance test (MMTT) was performed on the T2D/prediabetes participants to record their glucose and insulin responses to a standardized liquid meal containing predetermined amounts of macronutrients. The MMTT was conducted following an overnight fast of at least 8 hours. Participants were instructed to consume a standard liquid meal, Boost Original Nutritional Drink, in 10 minutes. Blood samples were collected at baseline (0 min), 15 minutes, and 30 minutes after the ingestion of Boost. The blood samples were collected in serum separator tubes and centrifuged immediately after collection to obtain serum for further analysis. **Figure S1** shows participants' glucose and insulin responses during the MMTT. The plots show distinct patterns in the glucose and insulin responses among participants with T2D, prediabetes, and without T2D/prediabetes.

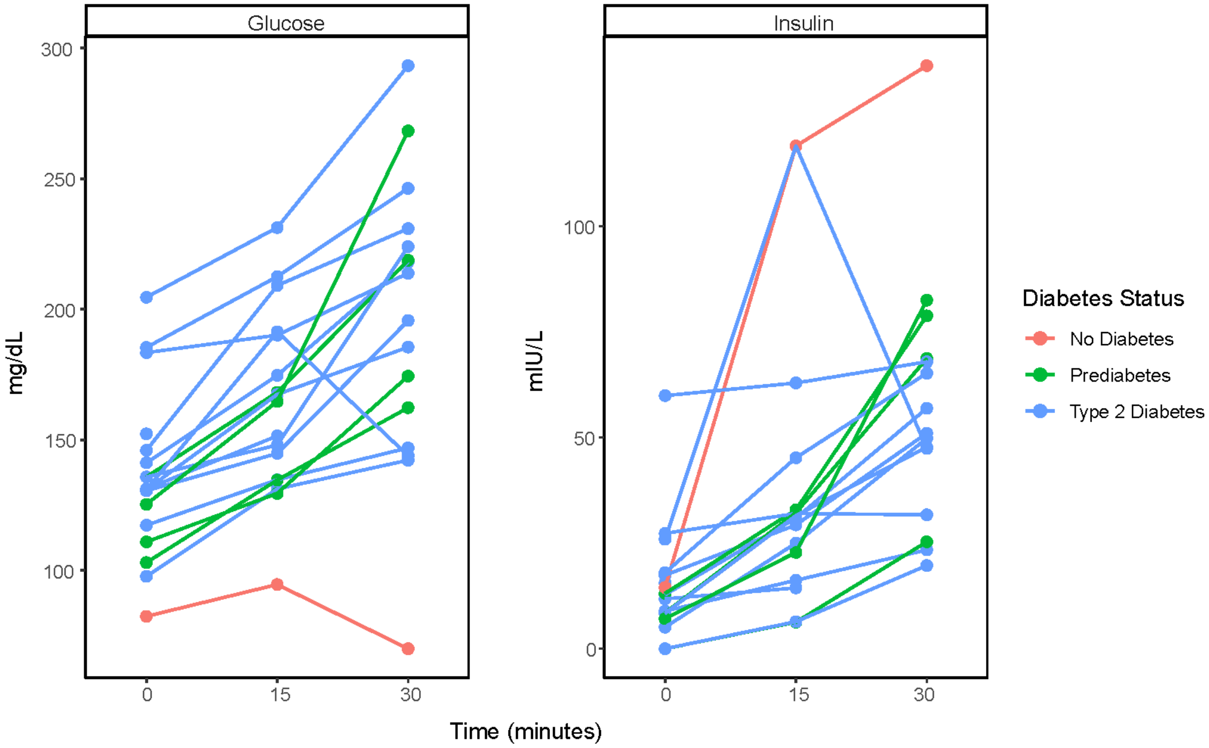


**Fig S1. Mixed meal tolerance test in participants with type 2 diabetes or prediabetes.** Plot illustrating the plasma glucose and insulin responses during a mixed meal tolerance test at 0, 15, and 30 minutes. The figure displays three diabetes status groups: orange–individual with no T2D/prediabetes (n = 1); green–individuals with prediabetes (n = 4); blue–individuals with T2D (n = 13). Mixed meal tolerance test data were not available for one participant with T2D.

| | Control (n = 76) | T2D/prediabetes (n = 18) | P-value |
|---|---|---|---|
| Age in years | 24.55 ± 4.48 | 65.3 ± 11.17 | <0.001 |
| Sex (%) | Female - 68% | Female – 37.5% | |
| | Male – 32% | Male – 62.5% | 0.04 |
| *AMY1* CN | 7.70 ± 2.8 | 6.37 ± 2.48 | 0.07 |
| SAA | 112.6 ± 86.5 | 159.25 ± 160.98 | 0.11 |

**Table S1. Table comparing individuals with T2D/prediabetes to those without T2D/prediabetes (controls).** This table shows the mean and standard deviation of the participant demographics. The healthy controls (n = 76) included individuals who submitted up to four saliva samples at different times of the day. The T2D/prediabetes group (n = 18) includes individuals who submitted up to two saliva samples.

Formula: lm(*AMY1* CN_ddPCR ~ *AMY1* CN_qPCR)
Coefficients:

| | Estimate | Std. Error | t value | Pr(>\|t\|) | |
|---|---|---|---|---|---|
| (Intercept) | 0.02093 | 0.18261 | 0.115 | 0.909 | |
| AMY1CN_q_PCR | 0.91711 | 0.02128 | 43.098 | <2e-16 | *** |

---
Signif. codes: 0 ‘***’ 0.001 ‘**’ 0.01 ‘*’ 0.05 ‘.’ 0.1 ‘ ’ 1

Residual standard error: 0.9192 on 208 degrees of freedom
Multiple R-squared: 0.8993, Adjusted R-squared: 0.8988
F-statistic: 1857 on 1 and 208 DF, p-value: < 2.2e-16

**Table S2. R output for linear regression to test the association between *AMY1* copy number values determined by qPCR and ddPCR.**

Formula: ICC(Dataframe [, c(AMY1_d_PCR, AMY1_q_PCR)], model = “twoway”, type = “consistency”, unit = “single”, conf.level = 0.95)

Subjects (sample#) = 210
Raters = 2
ICC(C,1) = 0.948

**Table S3. R output for the intraclass correlation coefficient (ICC) to assess consistency between qPCR and ddPCR.**

A. Formula: log(SAA) ~ time of saliva collection + (1 | participant_id)

Random effects:

| Groups | Name | Variance | Std.Dev. |
|---|---|---|---|
| participant_id | (Intercept) | 0.4919 | 0.7013 |
| Residual | | 0.1751 | 0.4184 |

Number of obs: 263, groups: participant_id, 76

Fixed effects:

| | Estimate | Std. Error | df | t value | Pr(>\|t\|) | |
|---|---|---|---|---|---|---|
| (Intercept) | 3.34757 | 0.22754 | 255.61340 | 14.712 | < 2e-16 | *** |
| Time of saliva collection | 0.08580 | 0.01684 | 220.37669 | 5.094 | 7.52e-07 | *** |

---
Signif. codes: 0 ‘***’ 0.001 ‘**’ 0.01 ‘*’ 0.05 ‘.’ 0.1 ‘ ’ 1

B. Saliva samples collected in the morning:

Formula: log(SAA) ~ time of saliva collection (morning samples) + (1 | participant_id)

Random effects:

| Groups | Name | Variance | Std.Dev. |
|---|---|---|---|
| participant_id | (Intercept) | 0.4859 | 0.6971 |
| Residual | | 0.1886 | 0.4343 |

Number of obs: 111, groups: participant_id, 51

Fixed effects:

| | Estimate | Std. Error | df | t value | Pr(>\|t\|) | |
|---|---|---|---|---|---|---|
| (Intercept) | 2.43678 | 0.63128 | 96.38994 | 3.860 | 0.000205 | *** |
| time_numeric | 0.17961 | 0.05938 | 93.08908 | 3.025 | 0.003216 | ** |

---
Signif. codes: 0 ‘***’ 0.001 ‘**’ 0.01 ‘*’ 0.05 ‘.’ 0.1 ‘ ’ 1

C. Saliva samples collected in the afternoon:

Formula: log(SAA) ~ time of saliva collection (afternoon samples) + (1 | participant_id)

Random effects:

| Groups | Name | Variance | Std.Dev. |
|---|---|---|---|
| participant_id | (Intercept) | 0.4648 | 0.6818 |
| Residual | | 0.1414 | 0.3760 |

Number of obs: 152, groups: participant_id, 63

Fixed effects:

| | Estimate | Std. Error | df | t value | Pr(>\|t\|) |
|---|---|---|---|---|---|
| (Intercept) | 3.41428 | 0.61089 | 130.42241 | 5.589 | 1.28e-07 *** |
| Time of saliva collection | 0.08503 | 0.04302 | 127.25596 | 1.976 | 0.0503 . |

---

Signif. codes: 0 ‘***’ 0.001 ‘**’ 0.01 ‘*’ 0.05 ‘.’ 0.1 ‘ ’ 1

**Table S4. A) R output for linear mixed regression testing association between SAA and the time of saliva collection during the day using samples collected in the morning and in the afternoon. B,C) R output for linear mixed models testing association between SAA and time of saliva sample collection in B) morning samples only and C) afternoon samples only.**

Formula: log(SAA) ~ time of saliva collection + time_of_day (morning or afternoon) + time of saliva collection * day_phase (morning or afternoon) + 1|participant

Random effects:

| Groups | Name | Variance | Std.Dev. |
|---|---|---|---|
| participant_id | (Intercept) | 0.4946 | 0.7033 |
| Residual | | 0.1733 | 0.4163 |

Number of obs: 263, groups: participant_id, 76

Fixed effects:

| | Estimate | Std. Error | df | t value | Pr(>\|t\|) |
|---|---|---|---|---|---|
| (Intercept) | 3.64910 | 0.58799 | 225.90375 | 6.206 | 2.57e-09 *** |
| Time | 0.06351 | 0.04137 | 219.49799 | 1.535 | 0.1262 |
| time_of_dayMorning | -1.16835 | 0.77551 | 215.72684 | -1.507 | 0.1334 |
| Time:time_of_dayMorning | 0.10693 | 0.06370 | 213.53953 | 1.679 | 0.0947. |

---

Signif. codes: 0 ‘***’ 0.001 ‘**’ 0.01 ‘*’ 0.05 ‘.’ 0.1 ‘ ’ 1

**Table S5. R output for linear mixed regression testing for the difference between rates of SAA change in the morning and the afternoon.**

Formula: log(SAA) ~ AMY1CN+1|participant_id

Random effects:

| Groups | Name | Variance | Std.Dev. |
|---|---|---|---|
| participant_id | (Intercept) | 0.4807 | 0.6933 |
| Residual | | 0.1997 | 0.4468 |

Number of obs: 300, groups: participant_id, 94

Fixed effects:

| | Estimate | Std. Error | df | t value | Pr(>\|t\|) |
|---|---|---|---|---|---|
| (Intercept) | 3.29795 | 0.22036 | 89.75471 | 14.966 | <2e-16 *** |
| AMY1 CN | 0.15266 | 0.02769 | 89.17525 | 5.512 | 3.4e-07 *** |

---

Signif. codes: 0 ‘***’ 0.001 ‘**’ 0.01 ‘*’ 0.05 ‘.’ 0.1 ‘ ’ 1

**Table S6. R output for linear mixed regression model testing for association between SAA and *AMY1* CN.**

```
Formula: log(SAA) ~ AMY1CN * health_status + time_of_day (morning vs afternoon) + Age +  Sex
+ (1 | participant_id)

Random effects:
 Groups         Name            Variance    Std.Dev.
 participant_id (Intercept)     0.4090      0.6395
 Residual                       0.1818      0.4264
Number of obs: 285, groups:  participant_id, 90

Fixed effects:
                              Estimate    Std. Error    df           t value   Pr(>|t|)
(Intercept)                   3.027695    0.340772      81.847562    8.885     1.24e-13 ***
AMY1CN                        0.131462    0.029137      79.265133    4.512     2.20e-05 ***
health_statust2d             -1.885951    0.585926      91.615135   -3.219     0.00178 **
time_of_dayMorning           -0.321545    0.067603     230.007694   -4.756     3.48e-06 ***
Age                           0.018423    0.007928      83.277524    2.324     0.02257 *
SexMale                      -0.018291    0.165042      83.655175   -0.111     0.91202
AMY1CN:health_statust2d       0.225122    0.079389      92.383203    2.836     0.00562 **
---
Signif. codes:  0 ‘***’ 0.001 ‘**’ 0.01 ‘*’ 0.05 ‘.’ 0.1 ‘ ’ 1
```

**Table S7. R output for linear mixed regression model assessing the effect of type 2 diabetes status on the association between *AMY1* copy number and salivary amylase activity.**
